# Exposure profiles in pregnant women from a birth cohort in a highly contaminated area of southern Italy

**DOI:** 10.1101/2022.11.09.22282107

**Authors:** Gaspare Drago, Silvia Ruggieri, Mario Sprovieri, Giulia Rizzo, Paolo Colombo, Cristina Giosuè, Enza Quinci, Anna Traina, Amalia Gastaldelli, Fabio Cibella, Simona Panunzi

**Affiliations:** National Research Council of Italy, Institute for Biomedical Research and Innovation, via Ugo La Malfa 153, 90146 Palermo, Italy; National Research Council of Italy, Institute of Anthropic Impacts and Sustainability in Marine Environment, via del Mare 3, 91021 Torretta Granitola, Trapani, Italy; National Research Council of Italy, Institute of Anthropic Impacts and Sustainability in Marine Environment, Lungomare Cristoforo Colombo 4521, 90149 Palermo, Italy; National Research Council of Italy, Institute of Clinical Physiology, Via Giuseppe Moruzzi 1, 56124 Pisa, Italy; National Research Council of Italy, Institute for System Analysis and Computer Science - BioMatLab, Via dei Taurini 19, 00168 Rome, Italy

**Keywords:** biomonitoring, persistent organic pollutants, mercury, food consumption, contaminated sites

## Abstract

Protecting the health of pregnant women from environmental stressors is crucial for reducing the burden of non-communicable diseases. In industrially contaminated sites, this action is particularly challenging due to the heterogeneous pollutant mixtures in environmental matrices. Aim of this study was to evaluate distribution patterns of mercury, hexacholobenzene and polychlorobyfenils in the serum of 161 pregnant women recruited in the framework of the NEHO cohort and living both inside and outside the National Priority Contaminated Site (NPCS) of Priolo. Food macro-categories were determined, and serum levels of contaminants were used to perform k-means cluster analysis and identify the role of food in pollutant transfer from the environment. Two groups of mothers with high and low measured pollutant levels were distinguished. Concentrations in mothers in the high-exposure cluster were at least twofold for all the evaluated pollutants (p<0.0001) and includes mothers living inside and outside NPCS, with predominance of individuals from the NPCS (p=0.045). Fish and vegetable consumption was higher in the high-exposure cluster (p=0.02). These findings suggest a direct link between marine sediments and soil contaminations, which in turn drives maternal exposure through the food chain. Such consideration appears poorly investigated in the context of contaminated sites.

**Highlights:**

- HCB, PCBs and Hg were measured in pregnant women from a highly contaminated area
- Pregnant women’s exposure profiles were obtained through k-means cluster analysis
- Distance of residence from emission sources only partially explains exposure levels
- Maternal age and food consumption appear to primarily drive contaminant levels

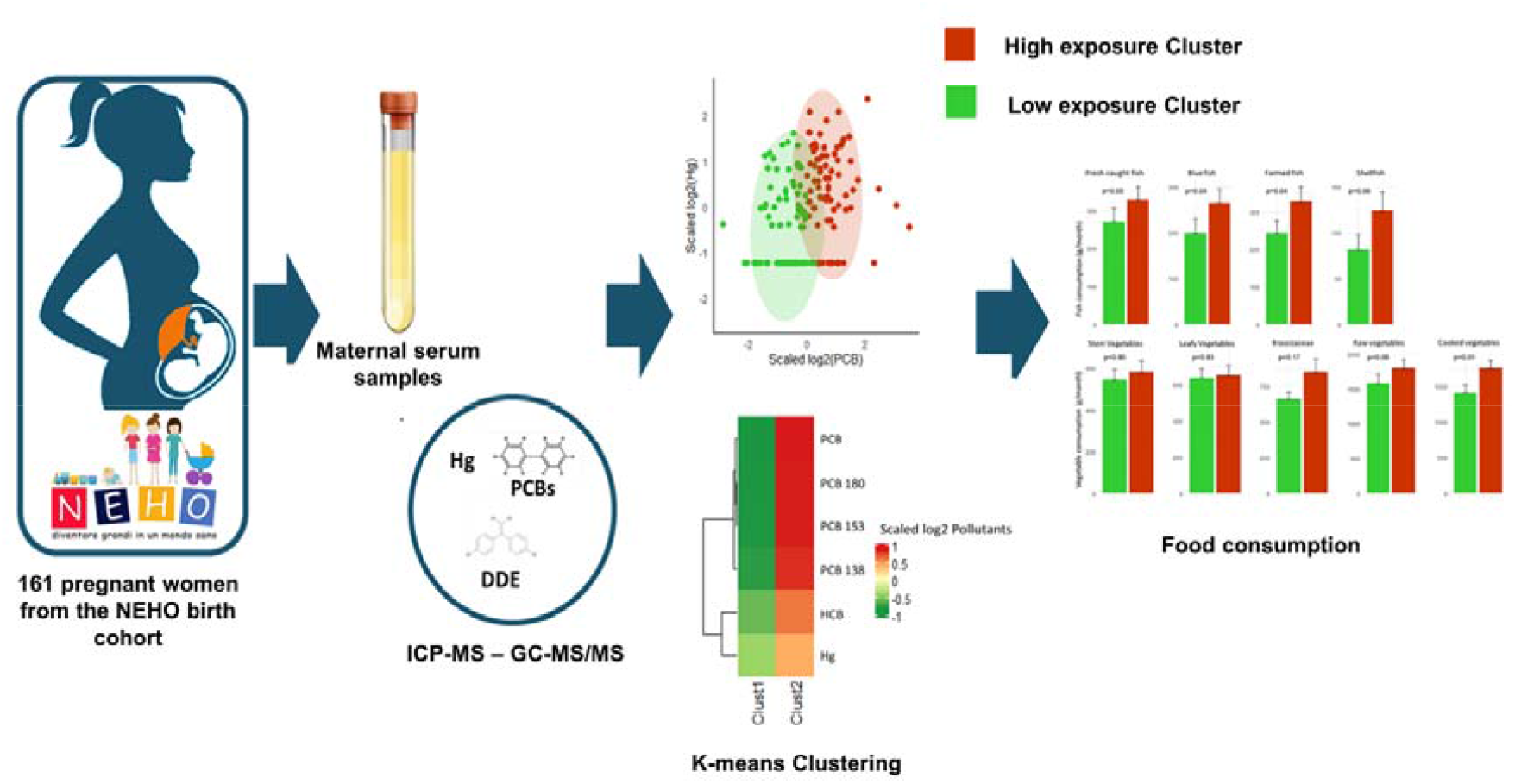

## 1. Introduction

A mounting body of evidence has convincingly linked the aetiology of several non-communicable diseases to environmental stressors. There is, therefore, an increasing need to prioritise strategies of environmental protection in order to significantly reduce the burden of chronic diseases (Sargis et al., 2019). In this perspective, the European Commission has published an ambitious agenda to challenge the effects of environmental degradation by 2050 (EU Commission, 2020). A parallel approach seeks to reduce the health consequences of environmental deterioration and points to identifying human exposure pathways in order to i) protect health by preventing/reducing the detrimental effects of hazardous agents, and ii) plan specific regulatory measures which can lead to improved environmental conditions (Bourguignon et al., 2018). Both these actions agree on recognising the prenatal and early postnatal time-interval as a period of life requiring major priority protection actions. In addition, both these lines of action are even more demanding in areas characterised by highly-complex industrial contamination where impacts on health, environmental pollution and socio-economic progress are deeply interconnected (Pirastu et al., 2013). People living near such contaminated areas are exposed to multiple and significant health threats. In fact, the simultaneous exposure to mixtures of several environmental pollutants – even at low concentrations – may produce a risk different from that produced by single contaminant exposure (van den Dries et al., 2021). Moreover, women of child-bearing age and developing foetuses are highly susceptible subgroups which are an absolute priority for safeguarding future generations (Martuzzi et al., 2004; Fuller et al., 2018; Drago et al., 2020). Exposure to contaminants that occurs during these ‘time windows’ may have long-term deleterious consequences during early infancy and may generate risks for major diseases later in life (Gluckman et al., 2010; Fleming et al., 2018).

In Italy, approximately 6 million people live within National Priority Contaminated Sites (NPCSs) where environmental degradation is a direct consequence and legacy of past industrialisation processes, often in the absence of effective and concrete environmental recovery policies (Pirastu et al., 2013). Priolo is a large industrialised marine-coastal area in southern Italy, hosting one of the largest European petrochemical plants. It covers an area of 550 km^2^ with a dense concentration of refineries, petrochemical and cement plants and waste dumps (Mudu et al., 2014). Within this area, Augusta Bay (the coastal-marine area of ∼25 km^2^ bordering the eastern boundary of the NPCS) was a main source of mercury (Hg) and other priority organic compounds discharged at sea until the 1970s as a by-product of a chloro-alkali plant and other petrochemical factories (Salvagio Manta et al., 2016; Denaro et al., 2020; Romano et al., 2021). In the same area, several studies have measured significant contamination by polychlorinated biphenyls (PCBs), hexachlorobenzene (HCB) and other classes of emerging pollutants detected in both environmental compartments and biota (Bellucci et al., 2012; Di Bella et al., 2020; Feo et al., 2020; Traina et al., 2021). Mercury and persistent organic pollutants (POPs) are long-lasting in the environment and bioaccumulate/biomagnify in tissues of living organisms. Foetal exposure to these chemicals during pregnancy can be a critical factor in a wide range of disorders later in life (Thayer et al., 2012; Gascon et al., 2013; Van Wijngaarden et al., 2013; Yorifuji et al., 2015; Saeedi Saravi et al., 2016). To understand the specific ways in which pregnant women may come into contact with such contaminants is a priority in order to verify the real extent of their health risks and quantify their exposure to toxic mixtures. In this light, human biomonitoring constitutes a powerful way to investigate how pollutant content in the organism varies following the dynamic of multiple exposure sources (Rappaport et al., 2012; Louro et al., 2019). This study is primarily aimed at evaluating the levels and distribution patterns of a few classes of pollutants – selected among some of those that specifically characterise the study area – in serum samples collected from pregnant women living in the Priolo area and enrolled in the Neonatal Environment and Health Outcomes (NEHO) cohort (Ruggieri et al., 2019). The study combines a wide range of information from a subset of questionnaires, with the aim of shedding light on possible causal links between lifestyle and detected serum contaminant concentrations. Therefore, we aimed to explore exposure profiles of 161 pregnant women from the Priolo area through the analysis of a mixture of pollutants, specifically Hg, HCB and three highly chlorinated PCB congeners with long biological half-lives (PCB138, PCB153 and PCB180). These specific contaminants had already been reported in several environmental compartments at relatively high concentration levels. This should offer crucial clues regarding the exposure mechanisms and pathways through which mixtures of environmental pollutants may affect such a critical population group.

## 2. Methods

### 2.1. Study sample

Between January 2018 and January 2020, the NEHO cohort enrolled 561 pregnant women living in the NPCS of Priolo and in Local Reference Areas (LRAs) located outside the NPCS boundaries but characterised by similar socio-demographic features (see Appendix ATable A1). For the present study, 161 women were randomly selected from the NEHO birth cohort^26^, 85 residing in the NPCS and 76 residing in LRAs: no significant difference was found between the selected sample and the whole birth cohort in Priolo area, with the exception of the numbers of previous pregnancies (Table A2, in Appndix A). Briefly, after reading a detailed information sheet, all the participants, during their last trimester of pregnancy, were required to sign a consent form confirming their understanding of the project’s aim. Detailed information was collected from the mothers using web-based questionnaires at enrolment and at 6, 12 and 24 months after delivery. Maternal blood samples were collected at enrolment, and serum was separated by centrifugation then temporarily stored at -20°C in each maternal unit before being transported on dry ice to the NEHO biobank for long-term storage at -80°C until the analysis was carried out.

The study was approved by the Ethics Committee “Catania 2” (July 11, 2017, No. 38/2017/CECT2). All procedures were conducted following the Declaration of Helsinki. The adopted protocol was compliant with the General Data Protection Regulation (UE 2016/679) and Italian data protection laws.

### 2.2. Questionnaire

Mothers enrolled in the NEHO cohort were asked to fill in different questionnaires. The “Baseline - first part” questionnaire provided information on maternal health and lifestyle during the gestational period. Some of the questions were aimed at retrieving information about the consumption habits of different types of food.

The maternal characteristics and socio environmental factors were age, body mass index before pregnancy (BMI), marital status, weeks of gestation and educational level, which originally was categorised into four levels: “Elementary school”, “Middle school”, “High school”, “Degree or higher qualification”. Here, the first two modalities were unified into the category “Secondary school or lower qualification”. Because the present study is also focused on the frequency of food consumption, the original questionnaire items collecting information about the consumption frequencies of the considered categories were modified as follows: “Never” was recorded in 0 days/month; “Once per month” was recorded in 1 day/month, “2/3 times per month” was recorded in 2.5 days/month; “Once per week” was recorded in 4 days/month; “2/3 times per week” was recorded in 10 days/month; “2/3 times per week” was recorded in 18 days/month; “Every day” was recorded in 30 days/month. A standard portion of each food was then considered in order to compute the total amount (in grams) consumed by each mother in one month. According to the National Recommended Energy and Nutrient Intake Levels (SINU, 2014), a standard fish portion corresponds to 150 g; 100 g is the standard portion for each type of meat; dairy products were expressed as a sum of yogurt 125 g, milk 125 g and fresh cheese 50 g; 200 g were used for eggs and vegetables, except for leafy and stem vegetables, for which a standard portion of 80 g was assumed. The quantity of vegetables consumed was computed as the sum of stem vegetables, leafy vegetables, Brassicaceae, raw and cooked vegetables.

### 2.3. Analytical procedure

Analyses of POPs (HCB and three congeners of PCBs 138, 153 and 180) in maternal serum were performed at the National Institute for Health and Welfare, Chemical Exposure Unit, Kuopio, Finland, with an Agilent 7000B gas chromatograph triple quadrupole mass spectrometer (GC-MS/MS). Ethanol and ^13^C-labelled internal standards were added to samples. Dichloromethane-hexane was added for extraction, followed by the addition of activated silica gel to bind the sample water and ethanol. The dichloromethane-hexane layer was poured into a solid phase extraction cartridge (SPE cartridge) containing 44% sulphuric acid silica, 10% silver nitrate impregnated silica and a mixture of sodium sulfate and silica. The lower semi-solid layer was extracted again with dichloromethane-hexane that was also poured into an SPE-cartridge. Elution of the SPE-cartridge was continued with dichloromethane-hexane, and the eluate was concentrated for GC-MS/MS. The quantification was performed by multiple reaction monitoring using an Agilent 7890A gas chromatograph/Agilent 7010 triple quadrupole mass spectrometer with DB-5MS UI column (J&W Scientific, 20 m, lD 0.18 mm, 0.18 μm). Reference materials for organic contaminants in human serum were analysed to estimate accuracy (SRM 1589a - National Institute of Standards and Technology, Gaithersburg, MD, USA). Recoveries ranged between 96% and 104% for each PCB and HCB analyte. Analytical precision was routinely better than 3% RSD%. Total serum triglycerides and cholesterol concentrations were assayed by certified spectrophotometric methods (Randox Laboratories, Crumlin, UK) at the Institute of Clinical Physiology of the National Research Council, Pisa, Italy. Total lipids were formulated according to the following equation (Covaci et al., 2006):

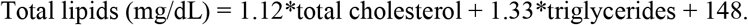

Lipid-standardised organochlorine concentrations were calculated from wet weight concentrations divided by total lipids.

Analyses of Hg were performed at the laboratory of LERES (Laboratoire d’Etude et de Recherche en Environnement et Santé) at the French School of Public Health - EHESP (Rennes, France), following the procedures described by Davies et al. (2021). The 161 serum samples were analysed by a plasma torch coupled with tandem mass spectrometer (ICP-MSMS 8800, Agilent Technologies) after a mineralisation step by adding nitric acid and heated with a heating block (Hotblock Pro, model SC-189, Environmental Express) at 83°C for 4 hours. Matrix effects correction was guaranteed through the use of internal standards (Sc, Ge, 77Se, Rh, Re and Ir). All internal standards were quantified in samples with less than 25% variation. Certified or internal control materials (measured additions) of blood and serum were added to the series (Utak level 1, Seronorm level 1) in order to guarantee the smooth running of the different stages and to cover the set of blood matrices. The results were validated since the concentrations of the controls were located within the limits of the control charts. This procedure was accredited by the French accreditation committee (CoFrac) in January 2020.

Concentrations below the limit of quantification (LOQ) were replaced by LOQ/2.

### 2.4. Statistics

Women were grouped according to their pollutant serum levels. A non-supervised K-means algorithm was used on the scaled logarithms to base 2. Concentrations were log-transformed to normalise their distribution and then scaled to homogenise their ranges. The Shapiro-Wilk test was used to determine whether the variables came from a normal distribution. The optimal number of clusters was estimated by computing both the Within Cluster Sums of Squares (WCSS), for the Elbow method, and the average silhouette.

The classification into clusters was used as a factor for testing associations with the NPCS and LRAs, as well as with other relevant qualitative variables using a Chi-Squared test or a Fisher exact test, when appropriate. The dependence of quantitative variables on the individuated clusters and the possible association between the clusters and the quantities of food consumed were assessed by means a Mann-Whitney U-test. A heatmap was used to graphically show the representative levels of blood pollutants in each cluster.

All the analyses were performed in R, version 4.1.3 (R software, 2020).

## 3. Results

### 3.1. Study sample characteristics

Table 1 reports a description of the enroled women with relevant demographic and socio-economic traits, separately by residence (in LRA or NPCS). Mean age (±SD) was 30.7±4.7 years, with no difference between the two groups (p=0.81). Similarly, BMI and the variable “weeks of gestation” emerged as not statistically different. The association between educational level and location was significant (p=0.02), highlighting a larger percentage of mothers with a higher educational level living in the NPCS. Marital status (married, never married/separated) was not significantly different (p=0.80), with the highest percentage of married women (65.0%) reflecting the distribution observed in the whole NEHO cohort (Ruggieri et al., 2021). Table 2 reports the pollutant concentrations in maternal serum of residents in LRAs and the NPCS (p values refer to the differences of contaminant concentrations between the two areas, tested by Mann-Whitney U-test). Pollutants in serum of women living in the highly contaminated area were significantly higher than those detected in samples from the reference areas, excluding Hg (p=0.402).

**Table 1.**
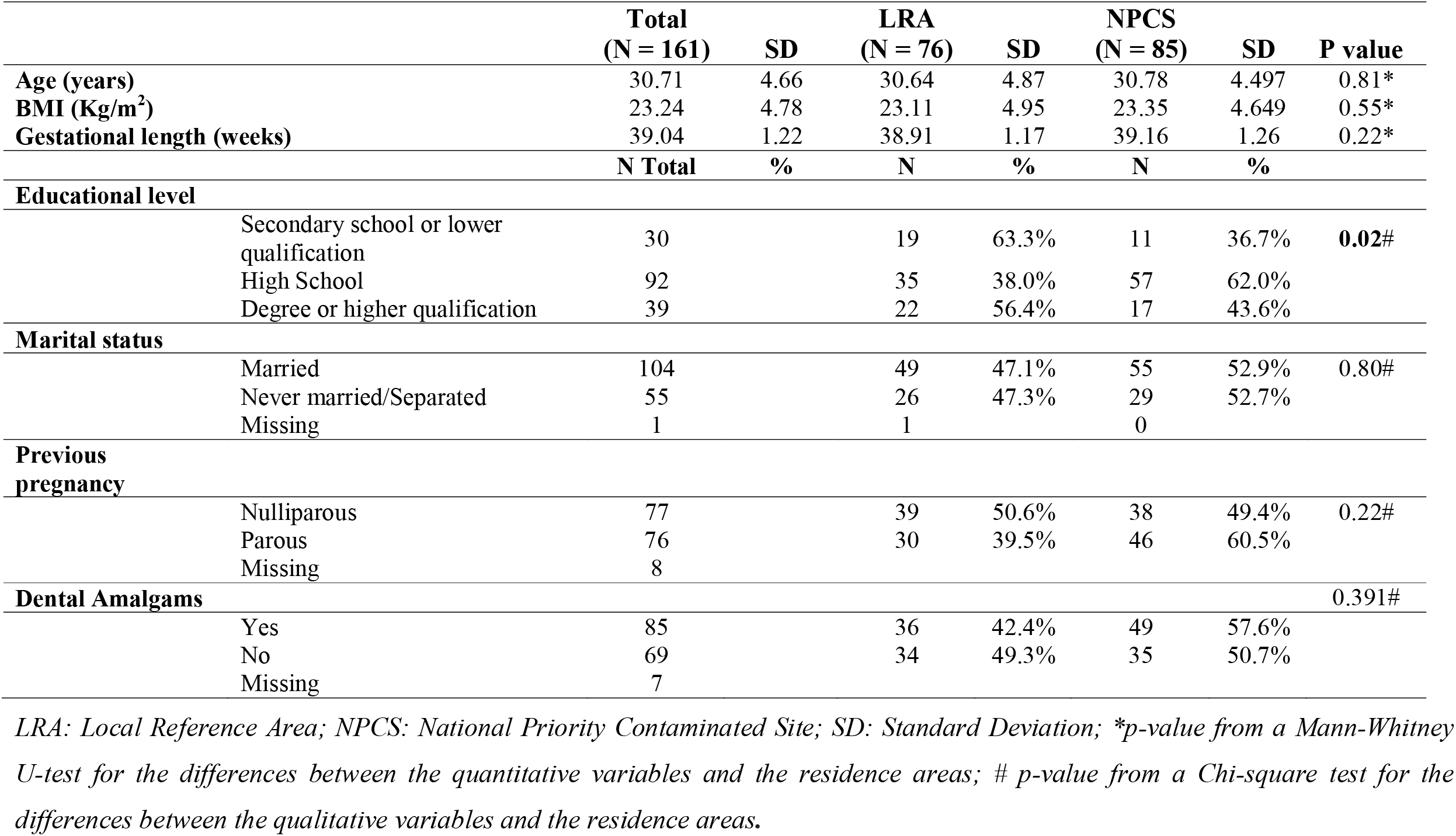
Socio-demographic characteristics of the study population.

|  |  | Total<br>(N = 161) | SD | LRA<br>(N = 76) | SD | NPCS<br>(N = 85) | SD | P value |
| --- | --- | --- | --- | --- | --- | --- | --- | --- |
| Age (years) |  | 30.71 | 4.66 | 30.64 | 4.87 | 30.78 | 4.497 | 0.81* |
| BMI (Kg/m<sup>2</sup>) |  | 23.24 | 4.78 | 23.11 | 4.95 | 23.35 | 4.649 | 0.55* |
| Gestational length (weeks) |  | 39.04 | 1.22 | 38.91 | 1.17 | 39.16 | 1.26 | 0.22* |
|  |  | N Total | % | N | % | N | % |  |
| Educational level |  |  |  |  |  |  |  |  |
|  | Secondary school or lower qualification | 30 |  | 19 | 63.3% | 11 | 36.7% | 0.02# |
|  | High School | 92 |  | 35 | 38.0% | 57 | 62.0% |  |
|  | Degree or higher qualification | 39 |  | 22 | 56.4% | 17 | 43.6% |  |
| Marital status |  |  |  |  |  |  |  |  |
|  | Married | 104 |  | 49 | 47.1% | 55 | 52.9% | 0.80# |
|  | Never married/Separated | 55 |  | 26 | 47.3% | 29 | 52.7% |  |
|  | Missing | 1 |  | 1 |  | 0 |  |  |
| Previous pregnancy |  |  |  |  |  |  |  |  |
|  | Nulliparous | 77 |  | 39 | 50.6% | 38 | 49.4% | 0.22# |
|  | Parous | 76 |  | 30 | 39.5% | 46 | 60.5% |  |
|  | Missing | 8 |  |  |  |  |  |  |
| Dental Amalgams |  |  |  |  |  |  |  | 0.391# |
|  | Yes | 85 |  | 36 | 42.4% | 49 | 57.6% |  |
|  | No | 69 |  | 34 | 49.3% | 35 | 50.7% |  |
|  | Missing | 7 |  |  |  |  |  |  |

*LRA: Local Reference Area; NPCS: National Priority Contaminated Site; SD: Standard Deviation; \*p-value from a Mann-Whitney U-test for the differences between the quantitative variables and the residence areas; # p-value from a Chi-square test for the differences between the qualitative variables and the residence areas.*

**Table 2.** Maternal serum contaminant levels. Serum levels of HCB and PCBs were normalized to total lipid content and reported in ng/g lipids.

| Pollutant | LOQ | N<br>(% ><br>LOQ) | Total (N=161) |  |  | LRA (N=76) |  |  | NPCS (N=85) |  |  | P*<br>value |
| --- | --- | --- | --- | --- | --- | --- | --- | --- | --- | --- | --- | --- |
|  |  |  | Mean<br>(SD) | Geom. Mean<br>(Min-Max) | Median<br>(IQR) | Mean<br>(SD) | Geom. Mean<br>(Min-Max) | Median<br>(IQR) | Mean<br>(SD) | Geom. Mean<br>(Min-Max) | Median<br>(IQR) |  |
| **Hg**<br>(µg/L) | 0.40 | 111<br>(68.9%) | 0.84<br>(0.75) | 0.59<br>(0.2-4.56) | 0.59<br>(0.2-1.21) | 0.80<br>(0.75) | 0.55<br>(0.2-4.56) | 0.57<br>(0.2-1.17) | 0.88<br>(0.76) | 0.62<br>(0.2-3.56) | 0.67<br>(0.2-1.22) | 0.402 |
| **HCB**<br>(ng/g) | 10.00<br>(ng/L) | 161<br>(100%) | 8.47<br>(6.19) | 7.39<br>(2.24-57.43) | 7.13<br>(5.55-9.79) | 7.89<br>(6.53) | 6.80<br>(2.24-57.43) | 6.78<br>(5.07-9.07) | 8.99<br>(5.85) | 7.97<br>(2.77-43.03) | 7.18<br>(6.05-10.43) | **0.041** |
| **PCB138**<br>(ng/g) | 5.00<br>(ng/L) | 161<br>(100%) | 9.88<br>(8.64) | 8.02<br>(1.59-84.03) | 7.78<br>(5.18-11.62) | 8.60<br>(6.78) | 7.07<br>(1.59-51.76) | 7.09<br>(4.76-10.77) | 11.02<br>(9.93) | 8.97<br>(2.55-84.03) | 8.97<br>(6.30-12.64) | **0.022** |
| **PCB153**<br>(ng/g) | 5.00<br>(ng/L) | 161<br>(100%) | 17.48<br>(15.67) | 13.97<br>(2.18-140.38) | 13.75<br>(9.17-21.41) | 15.23<br>(13.25) | 12.19<br>(2.18-103.45) | 11.80<br>(7.89-19.22) | 19.50<br>(17.39) | 15.77<br>(4.38-140.38) | 15.28<br>(10.58-22.53) | **0.016** |
| **PCB180**<br>(ng/g) | 5.00<br>(ng/L) | 161<br>(100%) | 12.90<br>(12.10) | 9.84<br>(1.07-91.27) | 9.89<br>(6.11-16.09) | 11.20<br>(10.73) | 8.45<br>(1.07-80.56) | 8.08<br>(5.08-13.79) | 14.42<br>(13.08) | 11.28<br>(2.96-91.27) | 11.33<br>(7.04-16.98) | **0.021** |
| **ΣPCB<sup>a</sup>**<br>(ng/g) | - | - | 40.26<br>(35.98) | 32.02<br>(4.84-315.67) | 31.52<br>(23.89-51.92) | 35.03<br>(30.52) | 27.88<br>(4.84-235.78) | 26.58<br>(17.62-44.10) | 44.93<br>(39.84) | 36.24<br>(9.88-315.67) | 34.69<br>(23.89-51.92) | **0.017** |

*LRA: Local Reference Area; NPCS: National Priority Contaminated Site; LOQ: limit of quantification; SD: Standard Deviation; IQR: Interquartile Range;*
*\*p-value from a Mann-Whitney U-test for the differences in pollutant levels between residence areas; ΣPCB<sup>a</sup>: Sum of PCB138, PCB153 and PCB180 congeners; concentrations below the LOQ were replaced by LOQ/2 before lipid adjustment.*

### 3.2 K-means clustering

From the k-means cluster analysis, both the Elbow and Silhouette methods identified 2 as the optimum number of clusters. Figure A1 (panels A and B in the Appendix A) shows the values of the two indices in correspondence with different Ks. The k-means procedure subdivided the sample into high-exposure (H-Exp) and low-exposure (L-Exp) groups. The heatmap in Figure 1 shows the average values of the pollutants in the two clusters (Panel A). Panel B shows how individuals were grouped into the two clusters based on the concentration levels of Hg and PCBs. Table 3 reports basic statistics of the measured pollutants from the two clusters. Table 4 shows the mothers’ distribution in the two clusters by their socio-economic traits. In particular, the association between clusters and area of residence (at risk vs local reference) was significant (p=0.045), with the largest percentage of women living in the NPCS belonging to the H-Exp cluster (47 of 77, 61%). In addition, individuals from this latter cluster were older than mothers with lower levels of contaminants (p<0.001) and with higher educational levels (p=0.018). The role of food consumption as driver of contamination to mothers was investigated by means of the Mann-Whitney U-test. Among the considered food categories (including meat, milk, eggs, fish and vegetables), consumption of fish and vegetables was significantly different in the two clusters (p=0.02 for both, Table 5). Specifically, Figure 2 (upper panel), shows the values of the average consumption of the categories of fish considered in the two k-means clusters (High and Low exposure levels). The p values from the Mann-Whitney U-test are also reported in the corresponding graphs. The average consumption of “Fresh caught fish”, “Blue fish” and “Farmed fish” resulted significantly different in the two clusters, with the women belonging to the H-Exp cluster consuming larger quantities of fish. Differently, “Shellfish” consumption was not significantly different between the two groups, also considering that shellfish are barely present in the diet of all the individuals studied. The average consumption of cooked vegetables was significantly higher in women belonging to the H-Exp cluster (p=0.014). Notably, while mothers of the H-Exp cluster preferentially consumed fish of local origin, no differences were found in terms of the provenance of vegetables (see Appendix A Table A3). However, in both cases, the consumption of products of local origin exceeded 70% of preferences.

**Figure 1.**
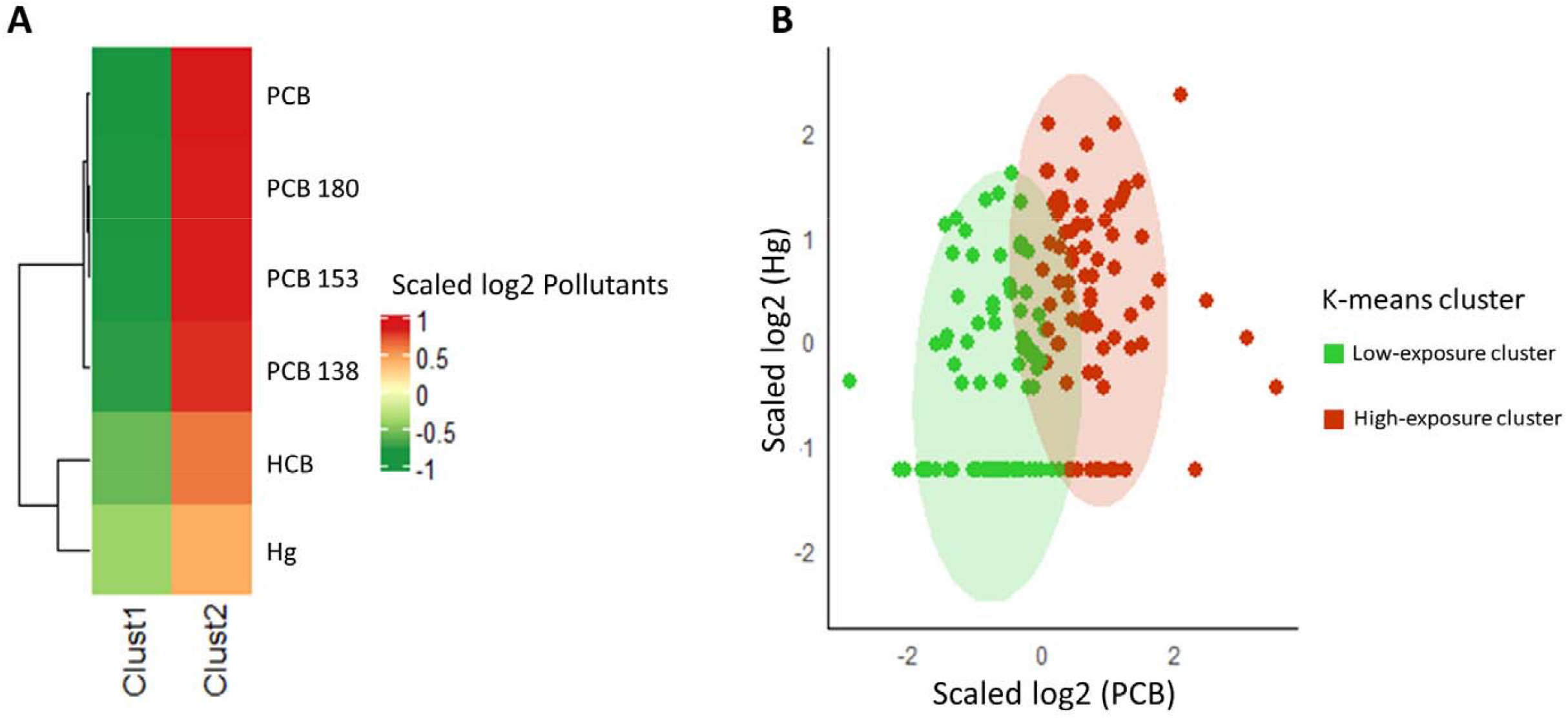
Panel A: Heatmap generated by k-means clustering analysis; Panel B: groups of individuals in the two clusters identified according to the levels of Hg and sum of the PCBs. Points aligned at the bottom of the figure represent the non-quantifiable values (i.e., values below the LOQ).

**Table 3.** Contaminant serum levels among the two identified clusters. Serum levels of HCB and PCBs were normalized to total lipid content and reported in ng/g lipids.

| Pollutant | Cluster 1<br>Low exposure |  |  | P<br>value<sup>§</sup> | Cluster 2<br>High exposure |  |  | P<br>value<sup>§</sup> | P value* |
| --- | --- | --- | --- | --- | --- | --- | --- | --- | --- |
|  | Total<br>(N=84) | LRA<br>(N=46) | NPCS<br>(N=38) |  | Total<br>(N=77) | LRA<br>(N=30) | NPCS<br>(N=47) |  |  |
|  | Median<br>(IQR) | Median<br>(IQR) | Median<br>(IQR) |  | Median<br>(IQR) | Median<br>(IQR) | Median<br>(IQR) |  |  |
| **Hg**<br>(µg/L) | 0.42<br>(0.20-0.74) | 0.40<br>(0.20-0.61) | 0.47<br>(0.20-0.84) | 0.509 | 0.96<br>(0.55-1.56) | 0.82<br>(0.55-1.51) | 1.04<br>(0.57-1.67) | 0.591 | < 0.0001 |
| **HCB**<br>(ng/g) | 5.83<br>(4.60-7.13) | 5.68<br>(4.34-7.60) | 5.93<br>(5.02-6.93) | 0.686 | 9.74<br>(7.32-11.88) | 8.82<br>(6.94-10.78) | 10.23<br>(7.49-12.37) | 0.159 | < 0.0001 |
| **PCB138**<br>(ng/g) | 5.34<br>(4.17-6.97) | 5.10<br>(3.72-6.96) | 6.06<br>(4.43-6.99) | 0.218 | 11.66<br>(9.57-15.73) | 11.46<br>(9.59-14.65) | 12.15<br>(9.57-17.21) | 0.409 | < 0.0001 |
| **PCB153**<br>(ng/g) | 9.41<br>(6.95-11.55) | 8.93<br>(5.83-11.44) | 9.69<br>(7.76-11.91) | 0.129 | 21.66<br>(16.99-28.04) | 21.66<br>(17.18-30.87) | 21.54<br>(16.81-25.12) | 0.616 | < 0.0001 |
| **PCB180**<br>(ng/g) | 6.46<br>(4.59-7.64) | 5.83<br>(3.86-7.62) | 6.96<br>(5.35-8.02) | 0.184 | 16.26<br>(12.50-21.21) | 15.34<br>(13.23-20.39) | 16.71<br>(12.46-23.12) | 0.794 | < 0.0001 |
| **ΣPCB<sup>a</sup>**<br>(ng/g) | 21.27<br>(15.90-26.25) | 20.02<br>(13.42-26.20) | 21.90<br>(17.40-27.27) | 0.153 | 49.68<br>(40.51-64.49) | 48.87<br>(39.96-62.69) | 50.75<br>(40.78-71.59) | 0.602 | < 0.0001 |

*IQR: Interquartile Range; \*p-value from a Mann-Whitney U-test for the differences between the two clusters; §p-value from a Mann-Whitney U-test for the differences between the residence areas within each cluster; ΣPCB<sup>a</sup>: Sum of PCB138, PCB153 and PCB180 congeners; concentrations below the LOQ were replaced by LOQ/2 before lipid adjustment.*

**Table 4.** Socio-demographic characteristics of the two clusters.

|  |  | **Total**<br>**(N=161)** | **SD** | **Cluster 1 (N=84)**<br>**Low exposure** | **SD** | **Cluster 2 (N=77)**<br>**High exposure** | **SD** | **P value** |
| --- | --- | --- | --- | --- | --- | --- | --- | --- |
| **Age (years)** |  | 31.3 | 4.7 | 29.3 | 4.3 | 33.5 | 4.1 | **<0.0001\*** |
| **BMI (Kg/m<sup>2</sup>)** |  | 23.2 | 4.8 | 23.5 | 5.4 | 22.9 | 4.0 | 0.834* |
| **Gestational length**<br>**(weeks)** |  | 39.0 | 1.2 | 39.1 | 1.2 | 40.0 | 1.3 | 0.650* |
|  |  | **N Total** |  | **N** | **%** | **N** | **%** |  |
| **Residence Area** |  |  |  |  |  |  |  | **0.045#** |
|  | NPCS | 85 |  | 38 | 44.7% | 47 | 55.3% |  |
|  | LRA | 76 |  | 46 | 60.5% | 30 | 39.5% |  |
| **Educational level** |  |  |  |  |  |  |  | **0.018#** |
|  | Secondary school or lower<br>qualification | 30 |  | 21 | 70.0% | 9 | 30.0% |  |
|  | High School | 92 |  | 49 | 53.3% | 43 | 46.7% |  |
|  | Degree or higher<br>qualification | 39 |  | 14 | 35.9% | 25 | 64.1% |  |
| **Marital status** |  |  |  |  |  |  |  | 0.388# |
|  | Married | 104 |  | 52 | 50.0% | 52 | 50.0% |  |
|  | Never married/Separated | 56 |  | 32 | 57.1% | 24 | 42.9% |  |
|  | Missing | 1 |  |  |  | 1 |  |  |
| **Previous pregnancy** |  |  |  |  |  |  |  | 0.572# |
|  | Nulliparous | 77 |  | 41 | 52.6% | 37 | 47.4% |  |
|  | Parous | 76 |  | 36 | 48.0% | 39 | 52.0% |  |
|  | Missing | 8 |  |  |  |  |  |  |
| **Dental Amalgams** |  |  |  |  |  |  |  | 0.323# |
|  | Yes | 85 |  | 40 | 47.1% | 45 | 52.9% |  |
|  | No | 69 |  | 38 | 55.1% | 31 | 44.9% |  |
|  | Missing | 7 |  |  |  |  |  |  |

*LRA: Local Reference Area; NPCS: National Priority Contaminated Site; SD: Standard Deviation; \*p-value from a Mann-Whitney U-test for the differences between the quantitative variables and the two clusters. # p-value from a Chi-square test for the differences between the qualitative variables and the two clusters.*

**Table 5.**
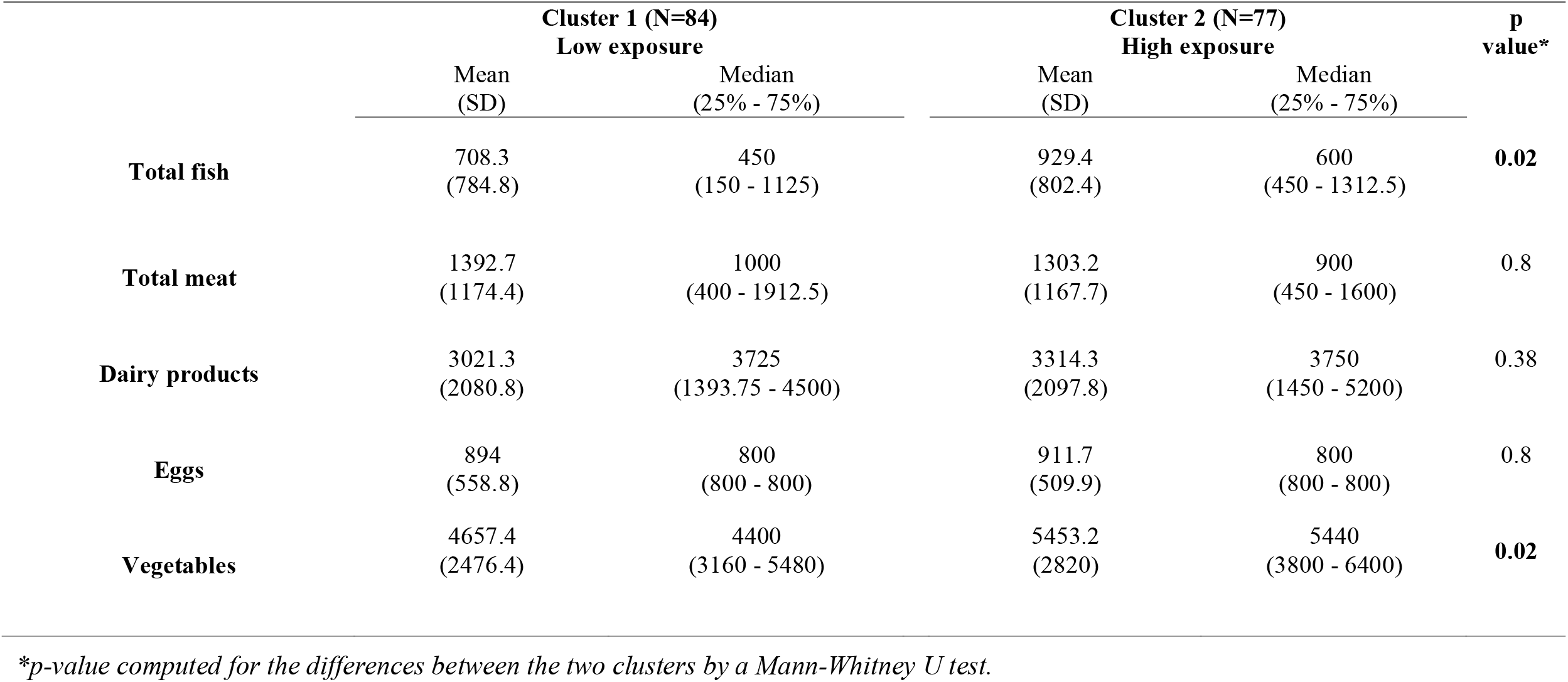
Consumption of food in g/month in the two clusters, identified by the k-means procedure.

|  | **Cluster 1 (N=84)**<br>**Low exposure** |  | **Cluster 2 (N=77)**<br>**High exposure** |  | **p**<br>**value\*** |
| --- | --- | --- | --- | --- | --- |
|  | Mean<br>(SD) | Median<br>(25% - 75%) | Mean<br>(SD) | Median<br>(25% - 75%) |  |
| **Total fish** | 708.3<br>(784.8) | 450<br>(150 - 1125) | 929.4<br>(802.4) | 600<br>(450 - 1312.5) | **0.02** |
| **Total meat** | 1392.7<br>(1174.4) | 1000<br>(400 - 1912.5) | 1303.2<br>(1167.7) | 900<br>(450 - 1600) | 0.8 |
| **Dairy products** | 3021.3<br>(2080.8) | 3725<br>(1393.75 - 4500) | 3314.3<br>(2097.8) | 3750<br>(1450 - 5200) | 0.38 |
| **Eggs** | 894<br>(558.8) | 800<br>(800 - 800) | 911.7<br>(509.9) | 800<br>(800 - 800) | 0.8 |
| **Vegetables** | 4657.4<br>(2476.4) | 4400<br>(3160 - 5480) | 5453.2<br>(2820) | 5440<br>(3800 - 6400) | **0.02** |

*\*p-value computed for the differences between the two clusters by a Mann-Whitney U test.*

**Figure 2.**
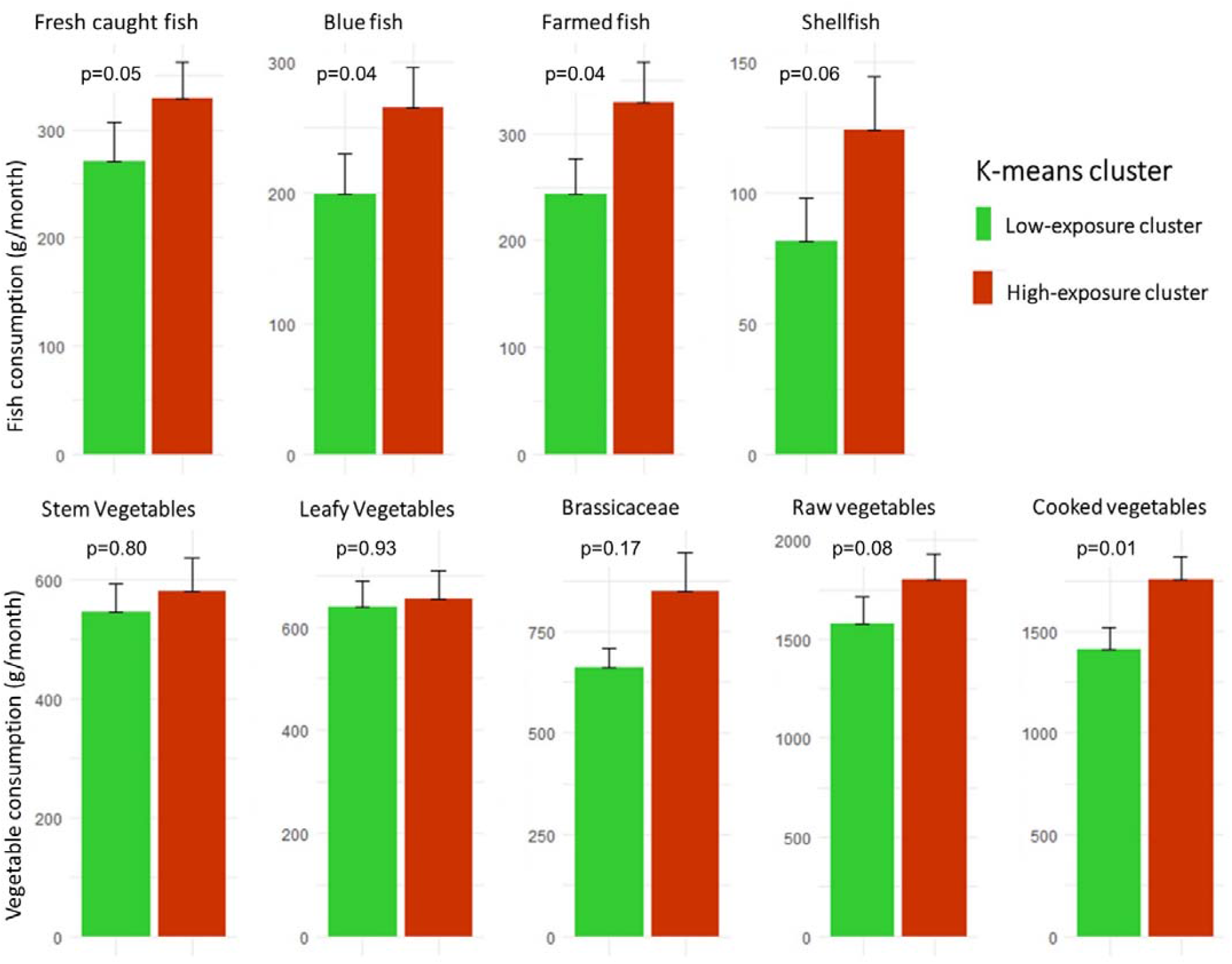
Average of fish (upper panel) and vegetable (lower panel) consumption in the two clusters. p-values from Mann-Whitney U-test.

Figure 3 shows the geographical distribution of mothers in the Priolo area according to the k-means clustering (H-Exp and L-Exp) and their area of residence (LRA vs NPCS). Residences of the mothers from the H-Exp cluster are shown in red, while those from the L-Exp cluster are in green. Moreover, circles and triangles discriminate between the mothers residing in the NPCS and LRA, respectively. In in the municipality of Augusta, within the NPCS, most of the mothers were associated with the H-Exp cluster (Figure 3, panel B).

**Figure 3.**
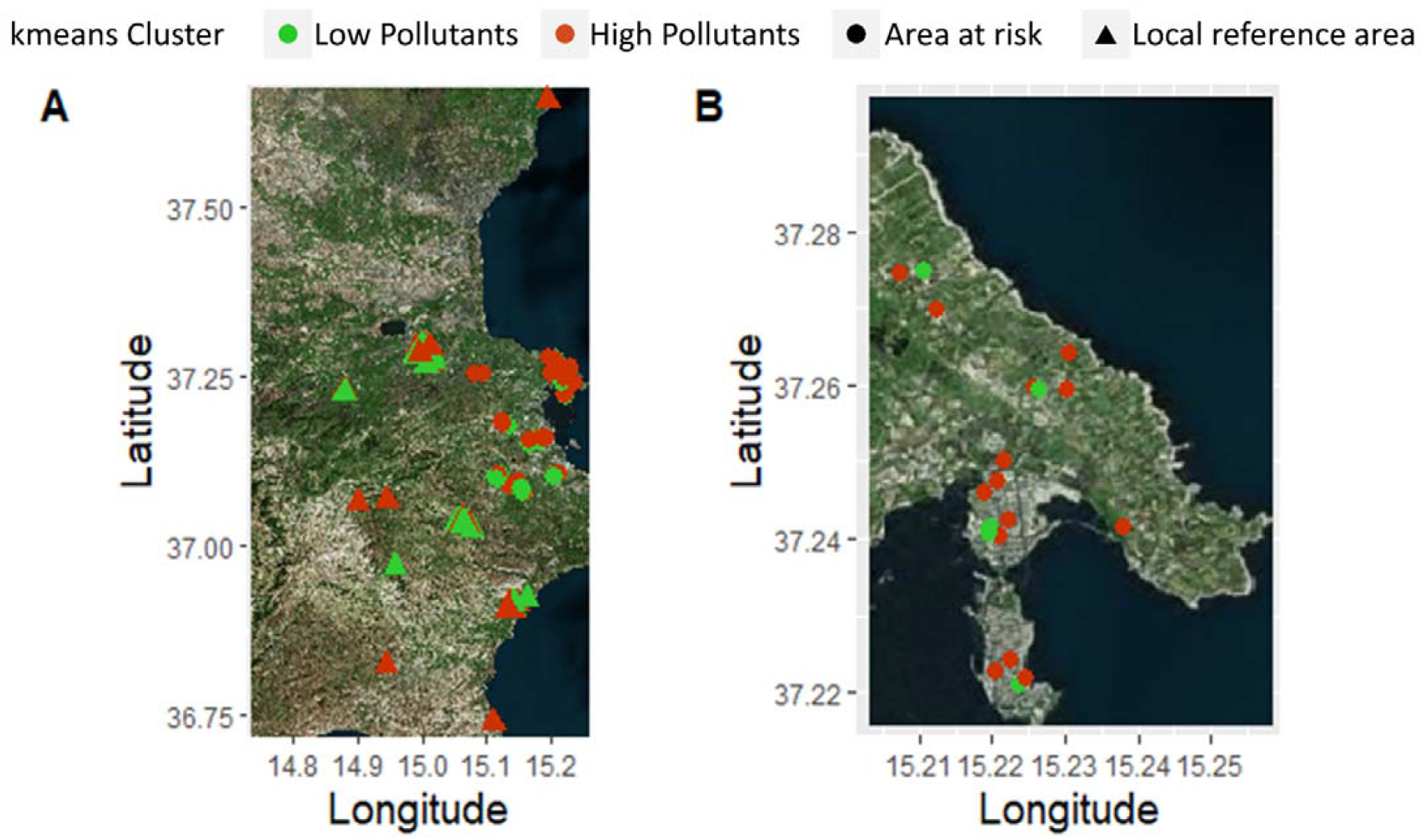
Geographical distribution of mothers in the Priolo area according to the k-means clustering (low vs high pollutant levels) and area of residence (NPCS vs LRA). Panel A refers to the entire study area. Panel B refers to the municipality of Augusta and to Augusta Bay.

## 4. Discussion

### 4.1 Comparison of biomonitoring results with worldwide databases

The results of the k-means cluster analysis applied to the 161 pregnant women suggest that (i) maternal residence only partially explains the higher levels of contaminants in cluster comparisons; (ii) the “higher exposure cluster” is characterised by a relatively higher consumption of local fish and vegetables; (iii) age and highest level of education represent significant risk factors for high serum concentration of pollutants.

In general, the concentrations of organochlorine (OC) compounds measured in this study were lower than those reported in pregnant women from other European countries such as Poland (Jaraczewska et al., 2006), Norway (Veyhe et al., 2015), the Netherlands (Soechitram et al., 2004), Denmark (Bjerregaard-Olesen et al., 2017) and Spain (Llop et al., 2010; Ibarluzea et al., 2011; García-Villarino et al., 2020). Similarly, a study conducted near the industrial area of Brescia, in northern Italy, reported higher OC levels in maternal serum (Bergonzi et al., 2009) than those found in this survey. Conversely, PCB concentrations were higher than those previously found in Japanese (Eguchi et al. 2022), Canadian (Fisher et al., 2016) and U.S. studies (Zota et al., 2013; Lyall et al., 2017; Ouidir et al., 2020). Moreover, the levels founded in our samples were very close to those reported by the multicentre European birth cohort study HELIX (Montazeri et al., 2019). based on data produced in six different European countries. To our knowledge, only two studies have reported concentrations of total Hg in the serum of pregnant women (Yau et al., 2014; Sekovanić et al., 2020). In particular, Yau et al. (2014) performed a case-control study to test the association between serum Hg levels and autism spectrum disorders, without documenting a meaningful association. The second study was conducted in Croatia where Sekovanic et al. (2020) analysed Hg in serum from mothers living both in continental and coastal areas. This latter study found significant differences in Hg concentration between the two areas but, in both cases, the levels of Hg reported in those studies were lower compared to our data. In 2011, Alimonti et al. (2012) piloted a wide biomonitoring survey in Italy, the PROBE study (PROgramme for Biomonitoring general population Exposure), which assessed the internal dosage of 20 metals in a representative sample of the Italian population. The levels of Hg in the Italian female population were lower on average than those found in our sample, in particular of women residing inside the NPCS (arithmetic mean = 0.70 μg/L and 0.88 μg/L respectively). This also reflects the outcomes of previous investigations in the Priolo area (Bonsignore et al., 2016; Di Bella et al., 2020) and indicates a crucial exposure of the local population to Hg.

### 4.2 Environmental contaminants and exposure pathways in the Priolo site

As shown in Table 1, no major differences in the socioeconomic variables were found between the two groups of enroled mothers, with the exception of their educational level, which appears higher in the NPCS. The comparison between the serum levels of selected contaminants in mothers from the NPCS and LRAs (Table 2) shows a significantly higher concentration of HCB and PCBs in the NPCS group, while the higher concentration of Hg was not statistically different (p = 0.402). Remarkably, unlike other studies (Bergdahl et al., 1998; Bedir Findik et al., 2016), in our samples, maternal Hg serum concentration was not associated with dental amalgam (p=0.60 – median and [IQR] of 0.60μg/L [<LOQ-1.20 μg/L] and 0.66 μg/L [<LOQ-1.22 μg/L] for mothers without and with dental amalgam, respectively). In addition, although higher concentrations of the same group of measured contaminants were found in other studies, the exposure levels required for endocrine disruption during pregnancy are reported to be extremely low (Zoeller et al., 2012; Vandenberg et al. 2014). However, synergistic or additive effects between pollutants have been increasingly documented (Kortenkamp et al., 2014; Magueresse-Battistoni et al., 2017; Longo et al., 2022). In light of this concern, we performed k-means cluster analysis aimed at identifying groups of mothers with different exposure levels. The two clusters show profiles of cumulative chemical exposure that might be associated with first-order indices of impact on health outcomes (Kalloo et al., 2018). Using a k-means clustering algorithm, we identified two distinct clusters of women based on serum contaminant concentrations. Specifically, pollutant levels in mothers from the H-Exp group were at least twofold higher in concentration with respect to the L-Exp group (Table 3). The median values of HCB and PCBs found in the H-Exp group exceeded the values reported in the above-mentioned work of Montazeri et al., which reported OC serum levels from six birth cohorts of different European countries (HCB median values = 9.74 ng/g vs 8.20; PCB138 = 11.66 vs 9.1; PCB153 = 21.66 vs 17.6 and PCB180 = 16.26 vs 10.4, respectively, for our sample and Montazeri et al.) ^44^.

As mentioned above, the H-Exp group contained 60% of mothers living within the NPCS and 40% residing in the control area. This emphasises that higher contaminant levels can be found not only in individuals living in the NPCS, but also in the LRAs, and that pollutants may be reasonably associated with common sources and pathways of contamination. Reasonably, local food and associated diets, reflecting the impact of environmental contamination, may represent a major pathway for transferring pollutants to humans, also for those populations living at a some distance from the emission site and primarily ‘linked’ to the same supply chains. Notably, the H-Exp group consisted mainly of older women, suggesting bioaccumulation effects for all the analysed pollutants (Philbert et al., 2000; Vizcaino et al., 2010; Ellsworth et al. 2018). This implies that developing foetuses were potentially exposed to higher contaminant levels in older pregnant women than in younger ones.

Such a ‘food hypothesis’ is also corroborated by evidence that mothers in the H-Exp group were characterised by significantly higher levels of fish and vegetable consumption. Mean total fish consumption in the H-Exp group (929.4 g/month) is in line with a previous study of fish consumption in 17 European birth cohorts (plus one American), which found an overall mean consumption of 1.5 times/week, corresponding to about 900 g per month (Stratakis et al., 2017). In the same study, the average consumption of fish for the Spanish birth cohort was 4.5 times/week, three times higher than those found in our cohort. These data could partially explain the higher levels of OCs found in their cohort than those found in our samples. The differences in fish consumption in the two clusters remain significant, even considering any individual fish category (Figure 2). Moreover, those in the cluster characterised by the highest exposure levels preferably consumed local fish (Table S3). Traina et al. (2021) reported a systematic correlation of PCBs (considering the same congeners), HCB and Hg between benthic commercial fish and marine sediments in Augusta Bay, thus demonstrating a robust fingerprinting of contamination pathways (Mudu et al., 2014). This definitively suggests a strong link between the highly polluted marine sediments of Augusta Bay that primarily drive benthic fish contamination, that, in turn, mirrors the high levels of (analogue) contaminants in pregnant women with diets characterised by preference for local fish. Thus, also in this case, the combination of Hg and selected OC compounds appears to represent a robust fingerprint for disentangling environmental impact mechanisms operating on the investigated group of pregnant women.

Vegetable consumption was significantly higher in the H-Exp cluster. No data for this class of food are available from the study area. Nonetheless, the achieved outcomes could suggest that soils, atmosphere and ground water contamination, as major environmental compartments potentially affecting pollution in vegetables, could play a critical role for population exposure. In line with what was found by Arrebola and colleagues in a Spanish adult population, vegetable consumption was significantly and positively associated with serum levels of PCBs (Arrebola et al., 2018). In addition, contamination of soils and plant crops near industrial sources of Hg has been previously documented (Li et al., 2017). This line of evidence needs specific pollutant analyses on vegetables from the study area to verify and conclusively constrain the proposed supposition.

Interestingly, we found that the H-Exp cluster was composed of a higher percentage of women with higher educational levels: this result is in agreement with our recent study (Ruggieri et al., 2022) showing that, in pregnant women, a higher educational stage enhances attention toward a “healthy” dietary pattern characterised by higher fish and vegetable consumption. Nevertheless, a similar diet, in a highly contaminated area, could produce a counterintuitive effect with a higher risk for exposure to environmental pollutants. With regard to fish consumption and its origin, our results confirm the data of the numerous studies carried out in the same areas on the risk of consuming local fish severely impacted by polluted sediments.

To our knowledge, this is the first biomonitoring study investigating serum levels of Hg and OCs in a sample of pregnant women residing in an NPCS. Despite the recruitment, performed on a voluntary basis, could have been influenced by a similar sociocultural level of the participants, joined by a common interest toward the health-related aspects of living in highly polluted areas, our findings highlight an urgent need to inform pregnant women living in highly contaminated areas about the risk arising from pollutants (Cori et al., 2019), as well as to suggest healthy lifestyle habits and diets. Remarkably, transfer routes of pollutants across the food chain and potentially reaching humans through daily diet appear priority areas of reseraches. This could inspire and support urgent and appropriate policies of interventions for mitigating environemntal impact on highly sensitive subgroups of population.

## Supporting information

Figure A1: Illustration of methods used to identify the optimal number of clusters. Table A1: List of municipalities included in the study and their l

## Data Availability

All data produced in the present study are available upon reasonable request to the authors

## Appendix A

Figure A1: Illustration of methods used to identify the optimal number of clusters.

Table A1: List of municipalities included in the study and their location with respect to industrial settlement.

Table A2: Comparison between the selected study sample and the whole birth cohort.

Table A3: Purchase origin of fish and vegetables and exposure cluster.

## Corresponding Author

*Silvia Ruggieri. Tel: +39 3662872999;

## Funding source

This work was developed within the CISAS International Centre of Advanced Study in Environment, Ecosystem and Human health, a multidisciplinary project on environment/health relationships funded by the Italian Ministry of Education, Universities and Research (MIUR) – CIPE resolution no. 105/2015 of 23 December 2015, grant number B62F15001070005.

## Acknowledgements

We wish to thank the colleagues involved in the *Piccolipiù* birth cohort for their support in defining questionnaires and the structure of the study. In particular, the present work has been carried out as a part of a scientific collaboration among the National Research Council of Italy - Institute for Biomedical Research and Innovation, Palermo, the Department of Epidemiology, Lazio - Regional Health System, Rome, and the Epidemiology Unit of the Anna Meyer Children’s University Hospital, Florence.

We would also like to thank the doctors and nurses working at the Umberto I Hospital in Syracuse and the General Hospital of Lentini, where the pregnant women were enrolled, for their fundamental support.

## Ethics approval

The NEHO study protocol has been approved by the Ethics Committee “Catania 2” for the NPCS of Priolo (11 July 2017, n. 38/2017/CECT2), and strictly followed the Declaration of Helsinki. Each participant read the information sheet and signed the informed consent. The participant information sheet is available at the NEHO website (www.neho.it). All the adopted procedures were compliant with the General Data Protection Regulation (UE 2016/679) and Italian laws concerning data protection.

## Author Statement

Gaspare Drago (G.D.), Silvia Ruggieri (S.R.), Mario Sprovieri (M.S.), Giulia Rizzo (G.R.), Paolo Colombo (P.C.), Cristina Giosuè (C.G.), Enza Quinci (E.Q.), Anna Traina (A.T.), Fabio Cibella (F.C.), Simona Panunzi (S.P.).

Conceptualisation, F.C., G.D., S.R. and S.P.; methodology, GD and S.P.; software, S.P. and G.R.; validation, F.C., G.D., S.R., and S.P.; formal analysis, S.P., G.D. and G.R.; investigation, S.R. and G.D.; resources, F.C.; data curation, G.D. and G.R.; writing—original draft preparation, G.D., S.R. and S.P.; writing—review and editing, F.C., S.P., G.D., P.C., M.S., E.Q., C.G., A.T. and visualisation, F.C., S.R., P.C., M.S., E.Q., C.G., A.T.; supervision, S.P. and F.C.; F.C. and M.S. coordinated the study and supervised the analysis of the data; project administration, F.C.; funding acquisition, F.C. All authors have read and agreed to the published version of the manuscript.

## Conflict of interest

The authors declare that they have no conflicts of interest.

## Notes

### Competing Interest Statement

The authors have declared no competing interest.

### Funding Statement

This study was funded by the Italian Ministry of Education, Universities and Research (MIUR) CIPE resolution no. 105/2015 of 23 December 2015, grant number B62F15001070005.

### Author Declarations

Ethics Committee Catania 2 of ARNAS Garibladi gave ethical approval for this work (11 July 2017, n. 38/2017/CECT2).

