## Supplementary material for "Exposure profiles in pregnant women from a birth cohort in a highly contaminated area of southern Italy": Figure A1: Illustration of methods used to identify the optimal number of clusters. Table A1: List of municipalities included in the study and their l

**Appendix A**

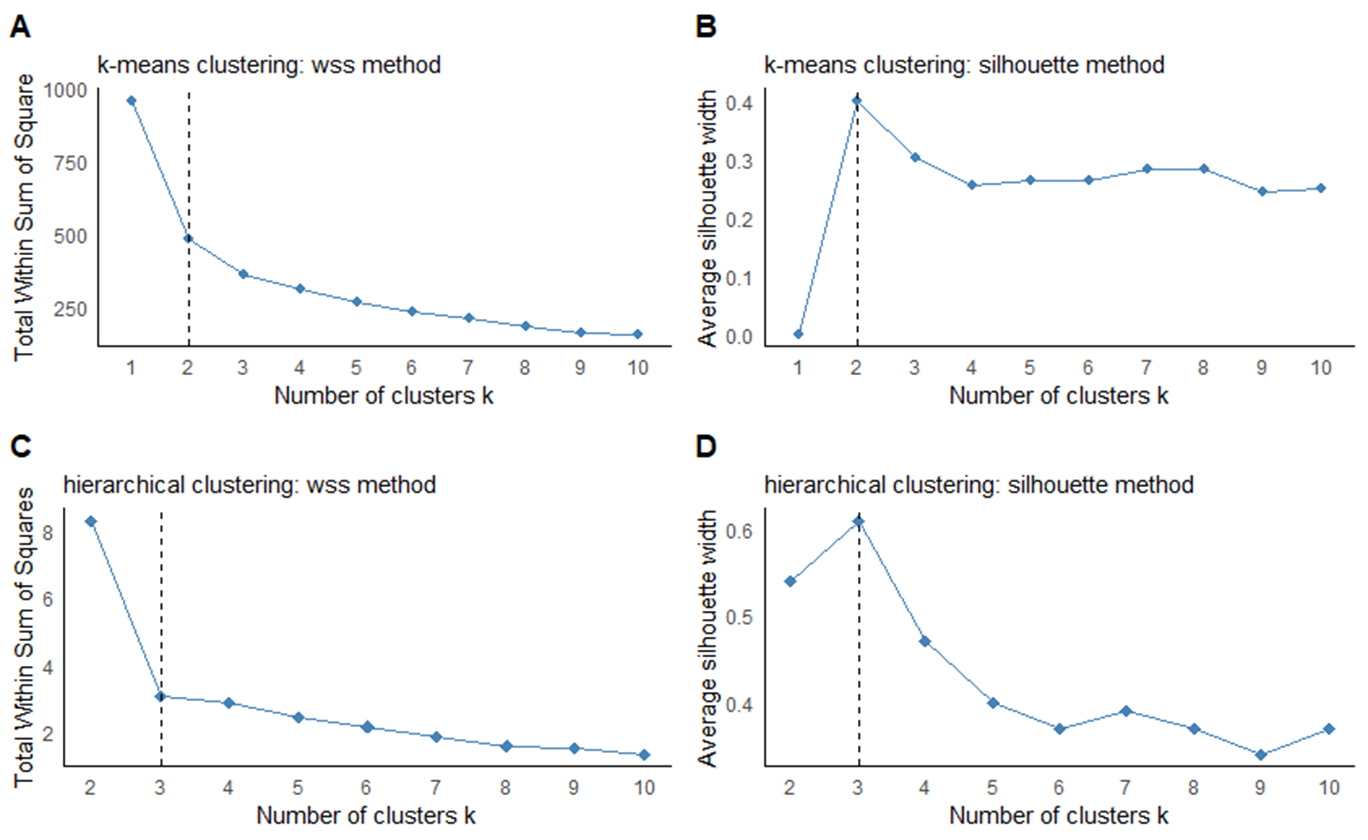

**Figure A1.** Elbow and Silhouette methods show the values of the two indices in correspondence to different Ks.

**Table A1.** Municipalities selected for the project in both study and local reference areas, with their respective distance from industrial settlement.

| **National Priority Contaminated Site** | **Municipalities in the study area** | **Municipalities in the reference area** | **Distance from industrial settlement (km)** |
| --- | --- | --- | --- |
| Priolo | Augusta  Priolo Gargallo  Melilli  Solarino  Floridia | Avola  Canicattini Bagni  Carlentini  Lentini  Noto  Pachino  Palazzolo Acreide  Rosolini  Francofonte  Palagonia  Scordia  Sortino  Vizzini | 27  19  22  22  45  59  51  57  49  61  50  25  63 |

**Table A2 –** Comparison of socio-demographic characteristics between the present study sample and the whole Priolo NEHO cohort.

|  | | | **Study sample (N = 161)** | | **NEHO Priolo cohort (N = 561)** | |  |
| --- | --- | --- | --- | --- | --- | --- | --- |
|  | | | **Mean** | **SD** | **Mean** | **SD** | **P value** |
| **Age (years)** | |  | 30.71 | 4.66 | 30.92 | 5.23 | 0.556* |
| **BMI (Kg/m^2^)** | |  | 23.24 | 4.78 | 23.60 | 4.85 | 0.341* |
| **Gestational length (weeks)** | |  | 39.04 | 1.22 | 39.41 | 1.23 | 0.754* |
|  | |  | **N Total** | **%** | **N** | **%** |  |
| **Educational level** |  | |  |  |  |  | 0.064# |
|  | Secondary school or lower qualification | | 30 | 18.63 % | 158 | 28.16% |  |
|  | High School | | 92 | 57.15 % | 277 | 49.38% |  |
|  | Degree or higher qualification | | 39 | 24.22 % | 124 | 22.10% |  |
|  | Missing | | - | - | 2 | 0.36% |  |
| **Marital status** |  | |  |  |  |  | 0.359# |
|  | Married | | 104 | 64.60 % | 341 | 60.80 % |  |
|  | Never married/Separated | | 55 | 34.16 % | 218 | 38.84 % |  |
|  | Missing | | 1 | 0.6 % | 2 | 0.36 % |  |
| **Previous pregnancy** |  | |  |  |  |  | 0.045# |
|  | Nulliparous | | 77 | 47.82 % | 259 | 46.17 % |  |
|  | Parous | | 76 | 47.21 % | 172 | 30.66 % |  |
|  | Missing | | 8 | 4.97 % | 130 | 23.17 % |  |
| **Dental Amalgams** |  | |  |  |  |  | 0.524# |
|  | Yes | | 85 | 52.80 % | 281 | 50.10 % |  |
|  | No | | 69 | 42.86 % | 199 | 35.47 % |  |
|  | Missing | | 7 | 4.34 % | 81 | 14.43 % |  |

*SD: Standard Deviation; *p-value from a Mann-Whitney U-test; # p-value from a Chi-square test*

**Table A3**. Percentages of purchase origin of fish and vegetables in relation to the k-means clusters.

|  |  | **Cluster 1**  **Low exposure** | **Cluster 2**  **High exposure** | **p value** |
| --- | --- | --- | --- | --- |
| Fresh caught fish | *Large retail chains* | 20.31 | 8.47 | 0.047 |
|  | *Local distribution* | 79.69 | 91.52 |  |
| Blue fish | *Large retail chains* | 20.75 | 4.08 | 0.016 |
|  | *Local distribution* | 79.24 | 95.92 |  |
| Farmed fish | *Large retail chains* | 21.31 | 7.02 | 0.036 |
|  | *Local distribution* | 78.69 | 92.98 |  |
| Shellfish | *Large retail chains* | 15.15 | 17.86 | 1 |
|  | *Local distribution* | 84.85 | 82.14 |  |
| Stem vegetables | *Large retail chains* | 26.25 | 25.68 | 0.961 |
|  | *Local distribution* | 73.75 | 74.32 |  |
| Leafy vegetables | *Large retail chains* | 21.52 | 22.08 | 0.945 |
|  | *Local distribution* | 78.48 | 77.92 |  |
| Brassicaceae | *Large retail chains* | 16.67 | 22.39 | 0.261 |
|  | *Local distribution* | 83.33 | 77.61 |  |
| Raw vegetables | *Large retail chains* | 23.68 | 25.00 | 0.377 |
|  | *Local distribution* | 76.32 | 75.00 |  |
| Cooked vegetables | *Large retail chains* | 28.57 | 23.88 | 0.663 |
|  | *Local distribution* | 71.43 | 76.12 |  |

*p-values from Mann-Whitney U-test*
